# Characterization of presumptive vancomycin-resistant enterococci recovered during infection control surveillance in Dallas, Texas

**DOI:** 10.1101/2020.10.09.20209569

**Authors:** Sara Ping, Nancy Mayorga-Reyes, Valerie J. Price, Michelle Onuoha, Pooja Bhardwaj, Marinelle Rodrigues, Jordan Owen, Dennise Palacios Araya, Ronda L. Akins, Kelli L. Palmer

## Abstract

*Enterococcus faecalis* and *E. faecium* are Gram-positive bacteria that normally inhabit the human gastrointestinal tract. They are also opportunistic pathogens and can cause nosocomial infection outbreaks. To prevent the spread of nosocomial infections, hospitals may rely on screening methods to identify patients colonized with multidrug-resistant organisms including vancomycin-resistant enterococci (VRE). Spectra VRE agar (Remel) contains vancomycin and other medium components that select for VRE and phenotypically differentiate between *faecalis* and *faecium* species by colony color. We obtained 66 de-identified rectal swab cultures on Spectra VRE agar that were obtained during routine patient admission surveillance at a Dallas, Texas hospital. We analyzed 90 presumptive VRE from 61 of the Spectra VRE agar cultures using molecular and culture methods. Using *ddl* typing, 55 were found to be *E. faecium* and 32 were found to be *E. faecalis*. While most of the *E. faecium* were positive for the *vanA* gene by PCR (52 of 55 strains), few of the *E. faecalis* were positive for either *vanA* or *vanB* (5 of 32 strains). The 27 *E. faecalis vanA*- and *vanB*-negative strains could not be recultured on Spectra VRE agar. Overall, we found that Spectra VRE agar performed robustly for the identification of vancomycin-resistant *E. faecium*, but presumptive false positives were obtained for vancomycin-resistant *E. faecalis*.

## Introduction

Vancomycin resistant enterococci (VRE), typically *Enterococcus faecium* and less commonly *E. faecalis*, are hospital-associated pathogens of significant public health concern. VRE are among the antibiotic-resistant pathogens identified by the United States Centers for Disease Control and Prevention as being serious threats to public health (1).

It is important to distinguish VRE from vancomycin-susceptible enterococci (VSE). VSE are normal colonizers of the human gastrointestinal tract (2). Like VRE, VSE can opportunistically cause infections, but more treatment options are available for infections with VSE. There are a number of surveillance methods that can be employed to identify patients colonized with VRE and to guide infection control practices in clinical settings. These methods typically involve culture-based screening of rectal swabs or fecal material on media containing vancomycin. One such medium is Spectra VRE agar (Remel). Spectra VRE agar contains 6 μg/mL vancomycin and proprietary chromogens that allow distinction between *E. faecium* and *E. faecalis* based on colony color.

In this study, we analyzed presumptive VRE obtained during routine patient admission screening for multidrug-resistant organisms (MDRO) at a Dallas, Texas, hospital. Our goals were to validate the isolates obtained from Spectra VRE agar cultures and to develop a strain collection to be used in future studies of enterococcal biology.

## Materials and Methods

### Culture methods and molecular biology procedures

Unless otherwise stated, enterococci were cultured at 37°C in brain heart infusion (BHI) broth or on BHI agar. Spectra VRE agar plates for independent testing of presumptive VRE isolates were purchased from Fisher Scientific. Genomic DNA was isolated using the DNeasy Blood and Tissue kit (Qiagen). Taq polymerase (New England Biolabs) was used for PCR. PCR products used for sequencing analysis were purified using the GeneJET PCR Purification kit (Thermo Fisher Scientific). Sequencing of PCR products was performed at the Massachusetts General Hospital CCBI Core facility.

### Collection of presumptive VRE

Presumptive VRE were collected at the Methodist Dallas Health System during routine MDRO surveillance. Surveillance culture screening was performed on patients admitted with at least one of these risk factors: hospitalization for ≥2 consecutive nights in the preceding 30 days, transferred from another medical facility, residence in a nursing home or extended/long term care facility, or the presence of decubitus ulcer or a draining wound. We obtained rectal swab cultures that were performed as a part of these routine admission surveillance procedures (UT-Dallas protocol number MR 14-448 and Methodist Health System protocol number P15MHS.0001A). The rectal swabs were plated on Spectra VRE agar. Instead of disposal after clinical surveillance was complete, Spectra VRE plates with visible growth were coded numerically, deidentified, and transferred to the University of Texas at Dallas for further analysis of presumptive VRE. One plate corresponded to one unique patient. Spectra VRE plates were collected from August 2015 through October 2015. At least one colony from each Spectra VRE plate was sub-cultured into BHI broth, incubated overnight, and stored at −80°C with 25% glycerol. If multiple colony phenotypes were observed from one Spectra VRE plate (for example, multiple different colony colors or morphologies), representative colonies of each phenotype were picked.

### Species determination and *van* typing

Primers used in this study are shown in **Table S1**. *ddl* primer sets used to identify *E. faecalis* and *E. faecium*, and the type of vancomycin resistance (*vanA* or *vanB*), were previously reported (3, 4). In the event that an isolate was neither *E. faecium* or *E. faecalis* by *ddl* PCR, the 16S rRNA gene was amplified using universal primers 8S and 1492R (5) and sequenced. At least 1300 non-ambiguous bases were used as query against the NCBI nr/nt database by megaBLAST. Hits with 100% query coverage and the highest percent sequence identity are reported. Boiled colonies were used as a template for PCR where possible; for some isolates, purified genomic DNA was used as a template.

### Vancomycin MIC

Vancomycin susceptibility was assessed for a subset of isolates using broth microdilution. Twofold serial dilutions of drug were made with BHI broth in a 96-well microtiter plate. An overnight culture was diluted to an OD_600nm_ of 0.001, and 5 μL were used to inoculate the wells of the plate. The MIC was recorded after 24 hours incubation at 37°C. Broth microdilution experiments were performed twice independently for each strain.

### Spectra VRE growth test

Select stocked isolates of interest were inoculated onto Spectra VRE agar to confirm whether growth occurred. Overnight cultures in BHI broth were diluted to an OD_600nm_ of 0.005 in 1-3 mL fresh BHI broth, then incubated at 37°C for 3.5 hours. 100 μL culture was spread plated onto Spectra VRE agar; for some isolates, 50 μL culture was spread over one half of the plate. A subset of strains were also inoculated on BHI agar supplemented with 6 μg/mL vancomycin. Plates were incubated for 24 hours at 37°C. *E. faecium* 1,231,410, a VanA-type VRE (6) and *E. faecalis* V583, a VanB-Type VRE (7, 8), were included as positive controls, yielding purple and light blue lawns, respectively.

### MLST and CRISPR-Cas analysis of select *E. faecalis* strains

The MLST database (https://pubmlst.org/efaecalis/) and sequencing of internal fragments of seven house-keeping genes were used as previously described (9) to determine the sequence type (ST) of 11 *E. faecalis* isolates. Novel STs were submitted to the database. PCR was used to screen 23 *E. faecalis* isolates for the presence of three previously identified *E. faecalis* CRISPR loci (CRISPR1-Cas, CRISPR2, and CRISPR3-Cas) (10, 11). Specifically, previously reported primer sets were used to screen for the presence of CRISPR1 *cas9* and CRISPR3 *cas9* (10). Strains with negative *cas9* results were then confirmed to lack the entire CRISPR-Cas loci by using previously reported primer sets that anneal outside the conserved chromosomal locations were the loci occur (10). CRISPR2 arrays were amplified and sequenced as previously described (10, 12). CRISPR2 spacer sequences were compared with a previously reported CRISPR2 spacer dictionary generated from 228 *E. faecalis* genomes (12).

## Results

### Initial analysis of Spectra VRE cultures

We obtained 66 Spectra VRE plates with visible bacterial growth, corresponding to 66 presumptive VRE-positive patients. Subsequently, 100 colonies of interest were identified and stocked for further analysis (Table S2). Based on colony color, 33 were presumptive *E. faecalis* VRE (light blue color), 64 were presumptive *E. faecium* VRE (navy blue or purple), and 3 colonies had atypical color (white) (Table S2).

### Confirmation of species prediction

Of the 100 stocked strains, 10 could not be revived from freezer stock (Table S2). Of these, 1 was a presumptive *E. faecalis* by colony color, and 9 were presumptive *E. faecium* by colony color. The inability to revive these isolates reduced our isolate number to 90 and our patient cohort size from 66 to 61.

Primer sets designed to amplify the *ddl* genes of *E. faecium* and *E. faecalis* (3, 4) were used to determine the species of the 90 presumptive VRE isolates. For strains where *ddl* PCR yielded negative results for both *E. faecalis* and *E. faecium* primer sets, the 16S rRNA gene was amplified and sequenced.

Of the 32 presumptive *E. faecalis* VRE that were recovered from freezer stock, 25 were confirmed to be *E. faecalis* by *ddl* PCR, and 7 were not confirmed by *E. faecalis ddl* PCR. Of these 7 strains, 6 were found to be *E. faecium* by *ddl* PCR, and 1 was found to be a presumptive *E. raffinosus* or *E. gilvus* by 16S rRNA gene analysis (99.78% sequence identity). In total, 78% of the colonies predicted to be *E. faecalis* based on Spectra VRE colony color were confirmed to be *E. faecalis* using *ddl* typing (Table S2).

Of the 55 presumptive *E. faecium* VRE that were recovered from freezer stock, 47 were confirmed to be *E. faecium* by *ddl* PCR, 1 was a mixed culture of *E. faecium* and *E. faecalis*, and 7 were not confirmed by *E. faecium ddl* PCR. Of these, all 7 were found to be *E. faecalis* by *ddl* PCR. In total, 85% of the colonies predicted to be *E. faecium* based on Spectra VRE colony color were confirmed to be *E. faecium* using *ddl* typing (Table S2).

Of the 3 atypical white colonies, two were found to be *E. faecium* by *ddl* PCR, and one was found to be *Staphylococcus epidermidis* by 16S rRNA gene analysis (99.85% sequence identity) (Table S2).

In summary, of the 90 presumptive VRE isolates analyzed using *ddl* typing, 55 were found to be *E. faecium*, 32 were found to be *E. faecalis*, 2 were found to be neither, and one was a mixed culture.

### Determination of vancomycin resistance type and re-testing for growth on Spectra VRE

Vancomycin resistance in enterococci is conferred by the synthesis of peptidoglycan precursors for which vancomycin has reduced binding affinity (13). Vancomycin resistance loci can be classified by the gene sequences for the D-alanine-D-lactate ligases, VanA and VanB, which are the most prevalent vancomycin resistance types for hospital-associated VRE (14). We screened our isolates for the presence of *vanA* and *vanB* using previously reported primer sets (3, 4).

Of the 55 *ddl*-confirmed *E. faecium*, 52 were positive for *vanA* and negative for *vanB*, and 3 were negative for both *vanA* and *vanB* (Table S3). Broth microdilution with vancomycin was performed for 14 of the 52 *vanA*-positive isolates to confirm their vancomycin resistance, and their vancomycin MICs were all ≥256 μg/mL, as expected. The 3 isolates that were negative for both *vanA* and *vanB* were assessed for growth on Spectra VRE agar, and they did not grow, indicating that the initial clinical cultures were false positives. Broth microdilution with vancomycin was performed for these three isolates. Two of the isolates had vancomycin MICs of 2 μg/mL. One isolate (163-2) had a vancomycin MIC of 512 μg/mL, despite being negative by PCR for both *vanA* and *vanB* and failing to be re-cultured on Spectra VRE. Further investigation is required to determine the genetic basis for the phenotypes observed for this strain.

Of the 32 *ddl*-confirmed *E. faecalis*, 4 were positive for *vanA* and negative for *vanB*, 1 was positive for *vanB* and negative for *vanA*, and the other 27 were negative for both *vanA* and *vanB* (Table S4). All *ddl*-confirmed *E. faecalis* were assessed for growth on Spectra VRE agar. Only the 5 *vanA*- or *vanB*-positive strains grew robustly on Spectra VRE. For the other 27 strains, no growth was observed, except that a single light blue colony was observed for each of isolates 107-2 and 110. Nine of these 27 *vanA*- and *vanB*-negative strains were also assessed for growth on BHI agar supplemented with 6 μg/mL vancomycin, and they failed to grow. Overall, these results suggest that the initial clinical cultures for 27 of the 32 *E. faecalis* strains were false positives on Spectra VRE. To follow up on these results, broth microdilution with vancomycin was performed for 5 of these isolates, and their vancomycin MICs were 2-4 μg/mL. Additional investigation is required to determine the mechanism for incongruence between initial clinical culture on Spectra VRE and subsequent molecular typing and Spectra VRE re-culture results.

Finally, the presumptive *E. raffinosus/E. gilvus* isolate was found to be *vanA*-positive and *vanB*-negative (Table S5). This isolate was assessed for growth on Spectra VRE, and it grew, forming light purple colonies. The broth microdilution vancomycin MIC for this strain is 128 μg/mL.

### MLST and CRISPR2 analysis of select *E. faecalis* isolates

We used MLST (9) and a previously published CRISPR-based typing method for *E. faecalis* (12) to assess phylogenetic relationships among a subset of the *E. faecalis* isolates. MLST uses sequence variation in 7 housekeeping loci to characterize phylogenetic relationships between strains. CRISPRs are hypervariable loci consisting of short (36 base pair) repeats interspersed by short (30 base pair) spacer sequences in *E. faecalis* (10, 11). Comparative analysis of CRISPR spacer sequences is an alternative method to MLST to analyze relationships between strains (15).

The sequence type (ST) of 11 *E. faecalis* isolates was determined, including 2 *vanA*-positive isolates and the single *vanB*-positive isolate. Eight STs were identified (Table 1). Five of the eleven isolates were novel STs (ST777, ST778, ST779), including the two *vanA* isolates (both ST779) and the *vanB* isolate (ST778). ST16 (2 isolates) and ST179 (one isolate) are single-locus variants of each other. Otherwise, all strains varied at ≥ 3 of the 7 loci relative to each other.

**Table 1.** MLST and CRISPR typing results.

| Isolate ID | *vanA/B*<sup>a</sup> | ST<sup>b</sup> | CRISPR2<sup>c</sup> | CRISPR-Cas<sup>d</sup> |
| --- | --- | --- | --- | --- |
| 2-1 | --- | 59 | 91-110-108-109-107 | Not present |
| 43-2 | --- | 64 | 85-83-86-82 | Not present |
| 59 | *vanA* | 779 | 92 | Not present |
| 80 | --- | ND | 85-83-86-82 | Not present |
| 81 | --- | ND | 12-67 | Not present |
| 94-1 | --- | 179 | 85-83-86-82 | Not present |
| 101-2 | --- | 6 | 12-67 | Not present |
| 106 | --- | ND | 85-83-86-82 | Not present |
| 107-2 | --- | ND | 1-5-38-70 | CRISPR1-Cas |
| 110 | --- | ND | 12-171(SNP)-170-89-169 | Not present |
| 119-1 | --- | 16 | 12-29-28-27-26-25-24-23-22-21 | CRISPR1-Cas |
| 122-2 | *vanA* | ND | 12-67 | Not present |
| 125-1 | --- | ND | 85-83-86-82 | Not present |
| 127-1 | --- | ND | 85-83-86-82 | Not present |
| 131-1 | *vanA* | 779 | 92 | Not present |
| 132 | --- | ND | 1-5-4-3-2 | Not present |
| 133-1 | --- | 777 | 85-a-b-c-d-e-f-g-h | Not present |
| 141-2 | *vanB* | 778 | 1-5-4-3-2 | Not present |
| 142-1 | --- | 777 | 85-i-b-j-e-f-g-h | Not present |
| 142-2 | --- | 16 | 12-29-28-27-26-25-24-23-22-21 | Not present |
| 143-1 | --- | ND | 85-83-86-82 | Not present |
| 153-1 | --- | ND | 92 | Not present |
| 152 | --- | ND | 85-83-86-82 | Not present |

<sup>a</sup>---, negative results for *vanA* and *vanB* PCR reactions.
<sup>b</sup>ND, not determined.
<sup>c</sup>Numbering scheme is from Hullahalli, et al, 2015. Letters indicate novel CRISPR spacers.
SNP, differs from spacer sequence 171 by a single nucleotide polymorphism.
<sup>d</sup>The presence or absence of CRISPR1-Cas and CRISPR3-Cas was evaluated for all isolates shown.

The CRISPR2 arrays of 23 *E. faecalis* isolates were amplified, sequenced, and analyzed (Table 1). We compared the CRISPR2 spacer sequences obtained here with our previous analysis of CRISPR2 loci in 228 *E. faecalis* genomes, wherein spacers of unique sequence were assigned unique numerical identifiers (12). We identified 12 CRISPR2 types among the 23 isolates. Only 2 of the 23 isolates analyzed here possessed novel spacer sequences relative to our previous study (denoted as letters in Table 1). Eight of the 23 isolates, corresponding to 8 patients, had identical CRISPR2 loci of spacers 85-83-86-82.

We also determined the occurrence of the previously identified CRISPR-Cas loci, CRISPR1-Cas and CRISPR3-Cas (10, 11), among the 23 *E. faecalis* analyzed for CRISPR2 typing (Table 1). Both of these systems can protect *E. faecalis* from the acquisition of mobile genetic elements that confer antibiotic resistance (16, 17). No isolates possessed CRISPR3-Cas. Two of the 23 isolates possessed CRISPR1-Cas (Table 1).

Two *E. faecalis* isolates from patient number 142 were analyzed by both MLST and CRISPR2 typing (Table 1). The isolates were of different STs and different CRISPR2 types, demonstrating co-colonization of the patient 142 gastrointestinal tract by genetically distinct *E. faecalis*.

## Conclusions

We report the collection and initial analysis of presumptive VRE obtained from hospitalized patients in Dallas, Texas. Our short-term goal (this study) was to validate the isolates obtained from Spectra VRE agar cultures. Our long-term goal is to use genome sequencing approaches to study the phylogeny and antibiotic resistance of these isolates.

We determined that most (52 of 55; 94.5%) of *ddl*-confirmed *E. faecium* from Spectra VRE rectal swab cultures were *vanA*-positive, while most (27 of 32; 84%) of *ddl*-confirmed *E. faecalis* were negative for both *vanA* and *vanB*. All of the *vanA*- and *vanB*-negative strains failed to be recultured on Spectra VRE, with the exceptions of a single colony observed for each of two *E. faecalis* strains. Conversely, control VRE strains and 6 *vanA*- and *vanB*-positive rectal swab strains were robustly cultured on Spectra VRE, as expected. There were key differences between use of the Spectra VRE agar in the clinical microbiology versus research labs. First, different batches of the agar were used. Second, the inoculation methods differed. In the clinical microbiology lab, rectal swabs (mixed cultures) were used to inoculate the agar, while in the research lab, high density pure cultures were spread on the agar. That said, of the 30 total isolates that failed to be recultured on Spectra VRE, we determined vancomycin MIC for 8 of them, and 7 had vancomycin MICs of 2-4 μg/mL. The concentration of vancomycin in Spectra VRE is 6 μg/mL, therefore it is expected that these strains should not grow on this medium. A final point is that in the clinical microbiology lab, the results of the entire agar culture are used to guide surveillance decisions, while in our research study, we selected only 1-3 single colonies per plate for analysis. Therefore, we cannot comment on whether the total Spectra VRE rectal swab culture was comprised of vancomycin-sensitive strains, or merely certain colonies. That said, we are not the first to observe false positives using Spectra VRE agar, particularly for *E. faecalis*. In a previous study, patient age was significantly correlated with false positive vancomycin-resistant *E. faecalis* results (18). For our strain collection, genome sequencing and more phenotypic assessments of vancomycin susceptibility and heterogeneity will be used in future studies to characterize these isolates further.

In conclusion, we present a collection of fecal enterococcal isolates from the Dallas, Texas, area. Most VRE identified via Spectra VRE agar were confirmed to be VanA-type *E. faecium*.

## Data Availability

All data are available in the paper.

## Acknowledgements

This project was supported by UT-Dallas start-up funds and R01AI116610 to K.P; a Louis Stokes Alliance for Minority Participation (LSAMP) scholarship to M.O., and the UT Dallas – Mexico Research Summer Program to N.M. We gratefully acknowledge Dr. Joslyn Pribble and the Microbiology lab at Methodist Health System, without whom this work would not be possible.

---

**Table S1.**
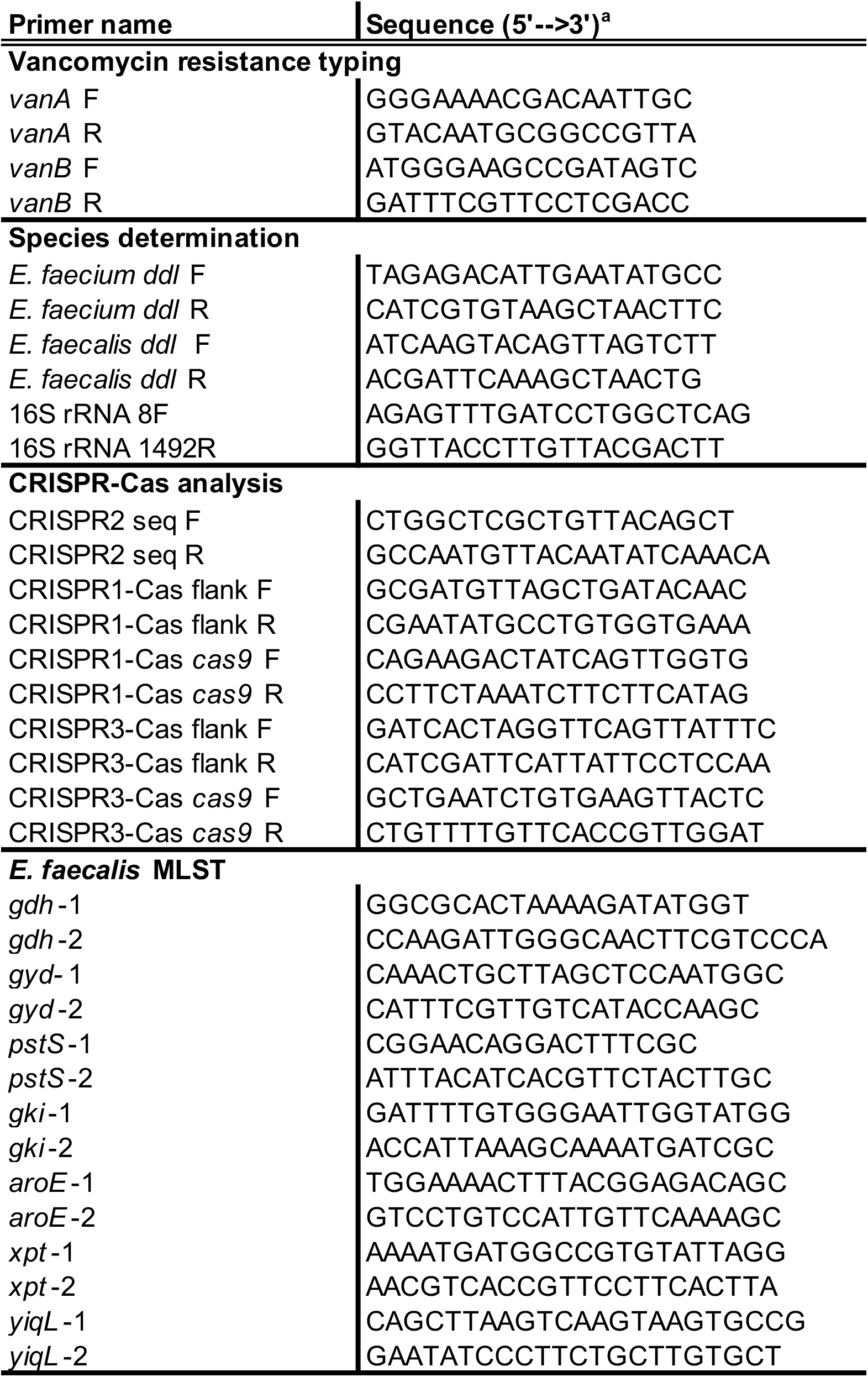
Primers used in this study. See materials and methods section for references for primers.

**Table S2.**
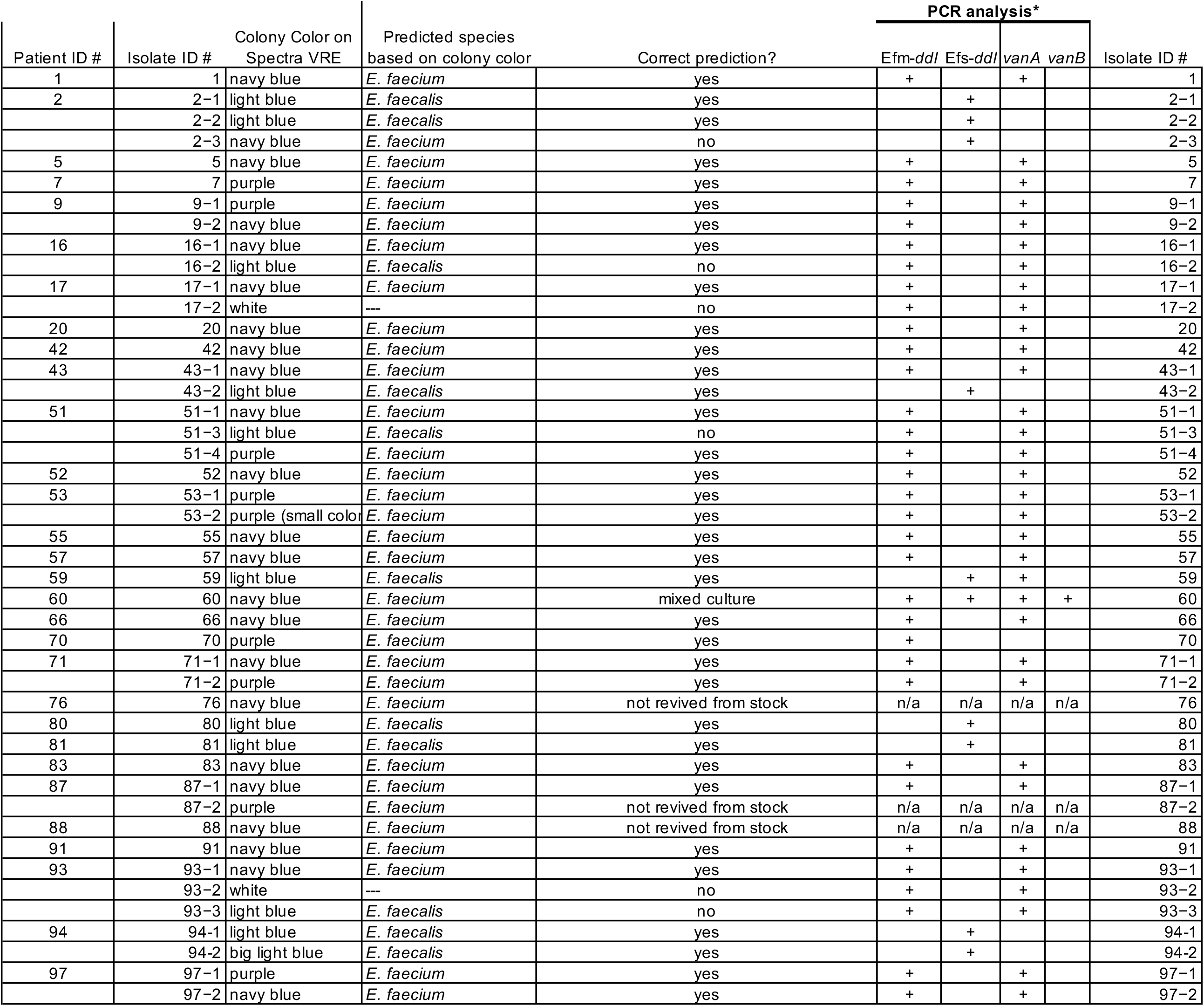
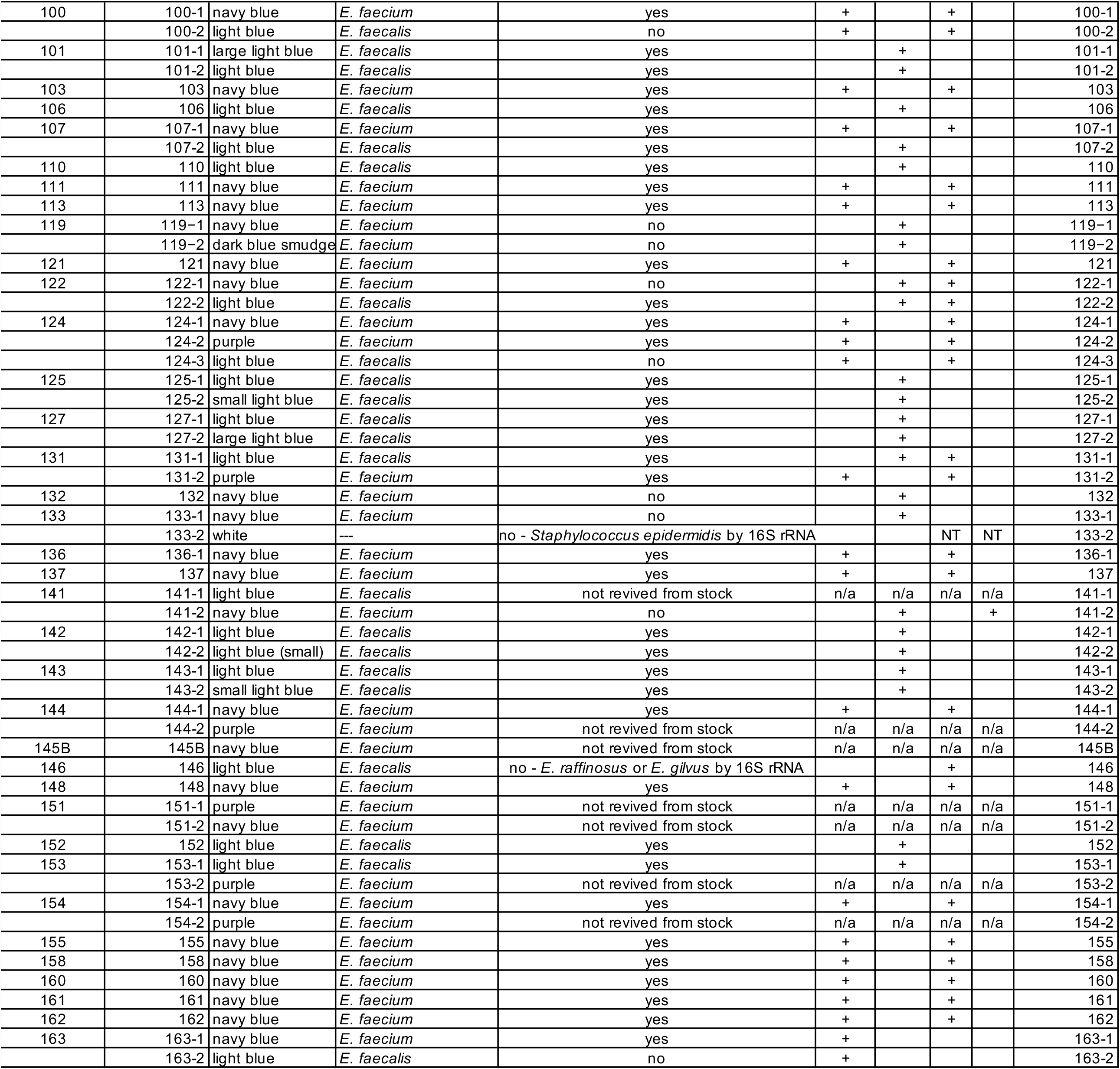
*ddl* typing data for presumptive VRE obtained from Spectra VRE plates. Notes: *For PCR analysis, an empty cell indicates a negative result for the PCR reaction. NT indicates Not Tested. n/a indicates that the PCR reaction was not performed because the strain could not be recovered from freezer stock.

**Table S3.** Data for *ddl* -confirmed *E. faecium* strains. Notes: *For PCR analysis, an empty cell indicates a negative result for the PCR reaction. NT indicates Not Tested. n/a indicates that the PCR reaction was not performed because the strain could not be recovered from freezer stock. ** For Spectra VRE re-testing, a blank cell indicates that the experiment was not performed for that strain. *** Two trials of broth microdilution were performed. If different MIC values were obtained across the two trials, both values are stated. A blank cell indicates that the experiment was not performed for that strain.

| Patient ID # | Isolate ID # | Colony Color on Spectra VRE | Predicted species based on colony color | Correct prediction? | PCR analysis* |  |  |  | Re-test for growth on Spectra VRE agar** | Vancomycin MIC by broth microdilution*** | Isolate ID # |
| --- | --- | --- | --- | --- | --- | --- | --- | --- | --- | --- | --- |
|  |  |  |  |  | Efm-*ddl* | Efs-*ddl* | *vanA* | *vanB* |  |  |  |
| 1 | 1 | navy blue | *E. faecium* | yes | + |  | + |  |  | 1024 | 1 |
| 5 | 5 | navy blue | *E. faecium* | yes | + |  | + |  |  | 2048 | 5 |
| 7 | 7 | purple | *E. faecium* | yes | + |  | + |  |  | 512 | 7 |
| 9 | 9-1 | purple | *E. faecium* | yes | + |  | + |  |  | 2048 | 9-1 |
|  | 9-2 | navy blue | *E. faecium* | yes | + |  | + |  |  |  | 9-2 |
| 16 | 16-1 | navy blue | *E. faecium* | yes | + |  | + |  |  | 1024 | 16-1 |
|  | 16-2 | light blue | *E. faecalis* | no | + |  | + |  |  |  | 16-2 |
| 17 | 17-1 | navy blue | *E. faecium* | yes | + |  | + |  |  | 512/1024 | 17-1 |
|  | 17-2 | white | --- | no | + |  | + |  |  |  | 17-2 |
| 20 | 20 | navy blue | *E. faecium* | yes | + |  | + |  |  | 1024 | 20 |
| 42 | 42 | navy blue | *E. faecium* | yes | + |  | + |  |  | 1024 | 42 |
| 43 | 43-1 | navy blue | *E. faecium* | yes | + |  | + |  |  | 2048 | 43-1 |
| 51 | 51-1 | navy blue | *E. faecium* | yes | + |  | + |  |  | 2048 | 51-1 |
|  | 51-3 | light blue | *E. faecalis* | no | + |  | + |  |  | 1024 | 51-3 |
|  | 51-4 | purple | *E. faecium* | yes | + |  | + |  |  | 1024 | 51-4 |
| 52 | 52 | navy blue | *E. faecium* | yes | + |  | + |  |  | 2048 | 52 |
| 53 | 53-1 | purple | *E. faecium* | yes | + |  | + |  |  | 256 | 53-1 |
|  | 53-2 | purple (small colony) | *E. faecium* | yes | + |  | + |  |  |  | 53-2 |
| 55 | 55 | navy blue | *E. faecium* | yes | + |  | + |  |  |  | 55 |
| 57 | 57 | navy blue | *E. faecium* | yes | + |  | + |  |  |  | 57 |
| 66 | 66 | navy blue | *E. faecium* | yes | + |  | + |  |  |  | 66 |
| 70 | 70 | purple | *E. faecium* | yes | + |  |  |  | No growth | 2 | 70 |
| 71 | 71-1 | navy blue | *E. faecium* | yes | + |  | + |  |  |  | 71-1 |
|  | 71-2 | purple | *E. faecium* | yes | + |  | + |  |  |  | 71-2 |
| 83 | 83 | navy blue | *E. faecium* | yes | + |  | + |  |  |  | 83 |
| 87 | 87-1 | navy blue | *E. faecium* | yes | + |  | + |  |  |  | 87-1 |
| 91 | 91 | navy blue | *E. faecium* | yes | + |  | + |  |  |  | 91 |
| 93 | 93-1 | navy blue | *E. faecium* | yes | + |  | + |  |  |  | 93-1 |
|  | 93-2 | white | --- | no | + |  | + |  |  |  | 93-2 |
|  | 93-3 | light blue | *E. faecalis* | no | + |  | + |  |  |  | 93-3 |
| 97 | 97-1 | purple | *E. faecium* | yes | + |  | + |  |  |  | 97-1 |
|  | 97-2 | navy blue | *E. faecium* | yes | + |  | + |  |  |  | 97-2 |
| 100 | 100-1 | navy blue | *E. faecium* | yes | + |  | + |  |  |  | 100-1 |
|  | 100-2 | light blue | *E. faecalis* | no | + |  | + |  |  |  | 100-2 |
| 103 | 103 | navy blue | *E. faecium* | yes | + |  | + |  |  |  | 103 |
| 107 | 107-1 | navy blue | *E. faecium* | yes | + |  | + |  |  |  | 107-1 |
| 111 | 111 | navy blue | *E. faecium* | yes | + |  | + |  |  |  | 111 |
| 113 | 113 | navy blue | *E. faecium* | yes | + |  | + |  |  |  | 113 |
| 121 | 121 | navy blue | *E. faecium* | yes | + |  | + |  |  |  | 121 |
| 124 | 124-1 | navy blue | *E. faecium* | yes | + |  | + |  |  |  | 124-1 |
|  | 124-2 | purple | *E. faecium* | yes | + |  | + |  |  |  | 124-2 |
|  | 124-3 | light blue | *E. faecalis* | no | + |  | + |  |  |  | 124-3 |
|  | 131-2 | purple | *E. faecium* | yes | + |  | + |  |  |  | 131-2 |
| 136 | 136-1 | navy blue | *E. faecium* | yes | + |  | + |  |  |  | 136-1 |
| 137 | 137 | navy blue | *E. faecium* | yes | + |  | + |  |  |  | 137 |
| 144 | 144-1 | navy blue | *E. faecium* | yes | + |  | + |  |  |  | 144-1 |
| 148 | 148 | navy blue | *E. faecium* | yes | + |  | + |  |  |  | 148 |
| 154 | 154-1 | navy blue | *E. faecium* | yes | + |  | + |  |  |  | 154-1 |
| 155 | 155 | navy blue | *E. faecium* | yes | + |  | + |  |  |  | 155 |
| 158 | 158 | navy blue | *E. faecium* | yes | + |  | + |  |  |  | 158 |
| 160 | 160 | navy blue | *E. faecium* | yes | + |  | + |  |  |  | 160 |
| 161 | 161 | navy blue | *E. faecium* | yes | + |  | + |  |  |  | 161 |
| 162 | 162 | navy blue | *E. faecium* | yes | + |  | + |  |  |  | 162 |
| 163 | 163-1 | navy blue | *E. faecium* | yes | + |  |  |  | No growth | 2 | 163-1 |
|  | 163-2 | light blue | *E. faecalis* | no | + |  |  |  | No growth | 512 | 163-2 |

**Table S4.**
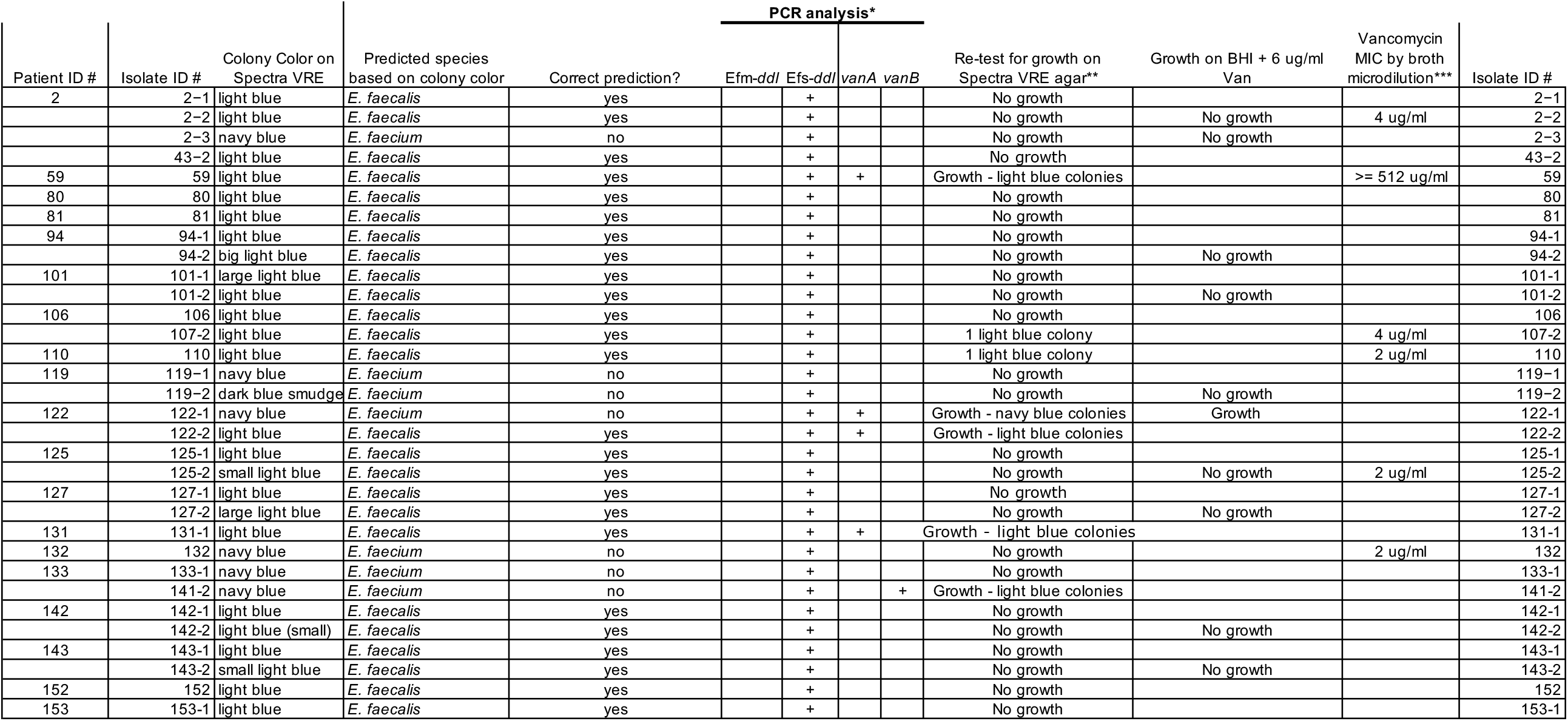
Data for *ddl* -confirmed *E. faecalis* strains. Notes: *For PCR analysis, an empty cell indicates a negative result for the PCR reaction. NT indicates Not Tested. n/a indicates that the PCR reaction was not performed because the strain could not be recovered from freezer stock. ** For Spectra VRE re-testing, a blank cell indicates that the experiment was not performed for that strain. *** Two trials of broth microdilution were performed. A blank cell indicates that the experiment was not performed for that strain.

**Table S5.**
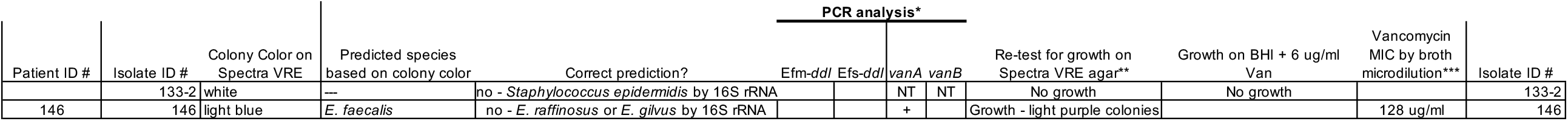
Data for species recovered other than *E. faecalis* or *E. faecium*. Notes: *For PCR analysis, an empty cell indicates a negative result for the PCR reaction. NT indicates Not Tested. n/a indicates that the PCR reaction was not performed because the strain could not be recovered from freezer stock. ** For Spectra VRE re-testing, a blank cell indicates that the experiment was not performed for that strain. *** Two trials of broth microdilution were performed. A blank cell indicates that the experiment was not performed for that strain.

